# ‘You’re still the same person, but you’re just seeing a bit less’: A qualitative study of the impact of vision impairment on mental wellbeing in adolescents with inherited eye disease

**DOI:** 10.64898/2026.07.28.26358577

**Authors:** Rachael F. Canavan, Marc S. Tibber, Ethan R. Tailor, Tessa M. Dekker, Michel Michaelides, Ngozi Oluonye, Dayyanah Sumodhee, Michael D. Crossland

**Affiliations:** UCL Institute of Ophthalmology, London, UK; Research Department of Clinical, Educational and Health Psychology, University College London, London, UK; Lived experience volunteer, Moorfields Eye-Young Persons’ Advisory Group; NIHR Moorfields Biomedical Research Centre, London, UK; Division of Psychology and Language Sciences, University College London, London, UK; Moorfields Eye Hospital NHS Foundation Trust, London, UK

## Abstract

**Objectives:** Mental wellbeing is lower in people with vision impairment (VI) but the mechanisms for this are not completely understood. Here we examined factors that limit and benefit wellbeing in children and young people with VI.

**Design:** Qualitative semi-structured interviews with children and young people with vision impairment and parents/carers of young people with VI. Interviews were recorded, transcribed and analysed by two researchers, working independently, using reflexive thematic analysis.

**Setting:** Low vision and genetic eye disease clinics in a tertiary eye hospital in London, England.

**Participants:** Young people aged 13-18 years with vision impairment (VI) caused by inherited macular disease and parents of young people with VI caused by inherited macular disease.

**Results:** Four overarching themes were developed: (1) *Living in the aftermath of diagnosis*, capturing participants’ internal experience of living with vision impairment from the time of diagnosis onward, including the psychological impact of negotiating the loss of vision and the challenge of accepting VI; (2) *Fighting the system*, reflecting how parents and CYP navigate both formal and informal support systems, highlighting the barriers they face; (3) *Being seen and being misunderstood*, encapsulating how VI influences the way that participants experience relationships, develop a sense of self and navigate their place within society; and (4) *Building a life with vision impairment*, centring on how participants function in everyday contexts whilst living with VI, alongside their aspirations for building their lives and living well in adulthood.

**Conclusions:** Vision impairment has emotional, social, and systemic consequences. Factors linking VI to reduced mental well-being included low acceptance of vision loss, low functional adaptation, reduced self-efficacy, effects of social stigma and discrimination.

**Strengths and limitations:**

- The study uses detailed interviews of wellbeing from two perspectives (young people and their parents/carers);
- Participants identified a broad range of positive and negative experiences which can be used to inform future interventions;
- All participants were under the care of a specialist eye clinic so they may not be fully representative of the broader population of people with vision impairment.

## INTRODUCTION

Living with vision impairment (VI) can impact negatively on the psychological and social wellbeing of adults[1–3] and of children and young people (CYP).[4,5] Although childhood vision impairment is relatively rare, around 1,000 children in England and Wales are registered as sight impaired or severely sight impaired each year[6] and globally around 1% of children have vision loss or blindness.[7]

In adults, mechanisms for reduced wellbeing in people with VI include activity limitation, loneliness, lower social support, lower self-efficacy, unemployment and having a later onset of VI.[2,8–13] Less is known about why wellbeing is lower in children and young people with VI, but factors such as social exclusion, low self-esteem and lack of independence have been proposed.[14–16] Although young people with VI are not thought to have smaller social networks than those without sight loss,[17] their participation in activities is often lower than their peers with no VI,[18] which may have important implications on their future loneliness.[19] Previous research has often investigated a heterogenous range of young people with VI, incorporating those with a variety of causes of VI (including cerebral VI and eye disease); those with no perception of light as well as those with some useful vision; those in specialist schools and those in mainstream education; and those who have VI from birth and those with acquired sight loss). The impact of these different modalities of VI on wellbeing is likely to be different although some shared mechanisms are likely to exist.

Emerging evidence has identified a range of interventions that may support well-being in this population. Psychological therapies such as cognitive behavioural therapy; rational emotive behaviour therapy; acceptance and commitment therapy (ACT); and emotional intelligence training have been associated with improvements in overall wellbeing of young people with VI.[20–24] Engaging in sport and exercise has also been linked to positive well-being and mental health outcomes,[22] as have activities such as music, art, reading, crafts, meditation and mindfulness.[25]

This study used qualitative research methods to explore the wellbeing of adolescents (aged 13-18) with lived experience of VI caused by inherited macular disease. Inherited macular diseases typically present in the second decade of life and cause progressive loss of central vision. All participants were in mainstream education and none had learning disability. Adolescents with lived experience and parents of children and young people with this condition were included, to explore their experiences and gain insight into both the impact of VI on well-being and their views on optimal support services and interventions.

## METHOD

This was a qualitative research study, exploring the association between VI and mental wellbeing, including an exploration of factors that enhance and limit wellbeing.

### Participant identification

Participants were recruited from a hospital low vision clinic in London, UK and a UK Based VI charity, and were invited to take part by author MDC. Study information was provided in an accessible format to young people and their parent or carer. Young people were included if they were aged between thirteen and eighteen years, had inherited macular disease and met the ICD–11 definition of VI caused by inherited eye disease (visual acuity poorer than 6/12 with both eyes open and/or a binocular visual field of less than 10°). Purposive sampling was used to ensure an age and gender mix of participants. Parent participants were over the age of eighteen (there was no upper age limit) and had at least one child who met the ICD–11 definition of VI from inherited macular disease. Participants had sufficient English language skills to complete an online interview, as assessed by the study team. No funding was available for interpreting services.

### Procedure

Participants completed one semi-structured remote interview with author RFC on an online platform (Zoom, version 6.1.10) between January and June 2025. Participants were at home. One CYP participant had a parent present during the interview but at all other interviews only the participant and researcher were present. Topic guides (supplemental material) were developed based on a review of literature [26] and findings from an earlier study of adults with VI caused by inherited macular disease (Canavan, in press). Open-ended questions were used to investigate the impact of VI, specifically focusing on the CYPs’ well-being. Participants were asked to share their perspectives on activities that may support CYPs’ well-being, as well as their opinions on existing support systems. To encourage richer responses, a consistent, yet flexible approach to data collection was employed, with pre-determined, non-directive prompts used. Participants were invited to share any additional relevant information at the end of the interview. Participants were given a £20 shopping voucher to thank them for their participation.

### Analysis

Interviews were recorded, anonymised, and transcribed verbatim by an external transcription service. Interviews were analysed using reflexive thematic analysis.[27] Reflexive thematic analysis was selected due to its flexibility and its independence from any single epistemological or theoretical framework.[28] Data were analysed by two independent researchers (authors RFC and MDC) on a line-by-line basis using NVivo (release 14.23.4; qsrinternational.com) following six phases: (1) familiarisation with the data; (2) generating initial codes; (3) constructing initial themes; (4) reviewing and developing themes; (5) refining, defining and naming themes; and (6) producing the final report.

Themes were created using an inductive *a posteriori* approach. Reflexive thematic analysis was conducted from a critical realist perspective, recognising that participants’ accounts represent mediated interpretations of reality, shaped by their social, cultural, and situational contexts.[29]

To avoid dehumanising participants’ lived experience, participants were given pseudonyms rather than numbers or codes when reporting.[30]

### Patient and public involvement statement

The study was developed in consultation with the Moorfields Eye-Young Person’s Advisory Group who discussed and provided feedback on the protocol (including advising on the topic guide and suggesting that we compensated participants for their time) reviewed participant information sheets and gave advice over the way in which the study was performed.

Author ET performed peer review of the manuscript and was asked to provide feedback on the accessibility and ease of engagement in the study, their satisfaction with their participation in the study, and their comments on the clarity of the manuscript.

### Researcher characteristics and reflexivity

Researchers’ position and contribution are essential and crucial aspects of reflexive thematic analysis, which requires the researcher to acknowledge, recognise, and reflect upon the impact of their subjectivity on the nature of the data collected.[31] Author RFC, a white British female PhD student with a background in psychology and mental health, acknowledges that her professional training and personal perspective may influence her research; therefore, ongoing reflexivity was utilised to recognise this. This researcher had no previous connection with the participants. Author MDC is a white British male optometrist with 25 years of experience in low vision clinics and research and a PhD in ophthalmology. He had met some of the participants in the clinic. The remaining authors contributed a rich reflective insight into the research from a broader, external position. Participants were aware that this research was conducted as part of RFC’s PhD study.

### Ethical statement

The study was performed in accordance with the Declaration of Helsinki and received ethical approval from the Health Research Authority (IRAS number 326182) following review by the London - City & East Research Ethics Committee. All participants provided written informed consent prior to taking part, including consent for their interviews to be video-recorded.

## FINDINGS

Six children and young people (mean age 15 years, range 13-18 years; three female) and twelve parents of CYP were recruited (9 female). An additional four young people and four parents/carers consented to taking part in the study but changed their mind before the interview took place and dropped out of the study without giving a reason.

Interviews lasted between 25 and 62 minutes, with a mean time of 40 minutes (standard deviation: 11.6).

Reflexive thematic analysis led to four overarching themes being developed (**figure 1**):

**Figure 1.**
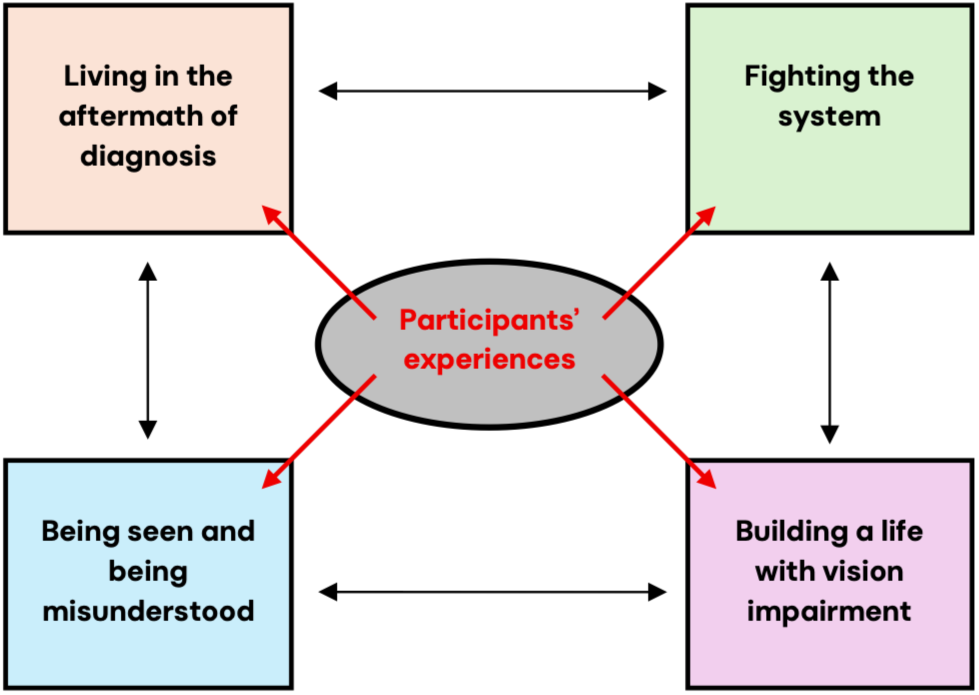
The four overarching themes developed through reflexive thematic analysis.

Theme 1. Living in the aftermath of diagnosis.

Theme 2. Fighting the system.

Theme 3. Being seen and being misunderstood.

Theme 4. Building a life with vision impairment.

These themes encapsulate the emotional, relational, structural and future-oriented dimensions of participants’ experiences, including the challenges encountered and the strategies developed in response. The themes are interconnected and highlight how psychological well-being, social relationships, support and future aspirations connect to shape participants’ experiences of living with VI.

### Theme 1. Living in the aftermath of diagnosis

This theme captured participants’ internal experience of living with VI from the time of diagnosis onward including the psychological impact of negotiating the loss of vision and the challenges of accepting VI.

Participants shared their initial emotional responses to the diagnosis. Parent participants described how stressful it was when their child was diagnosed and how they were trying to navigate their own emotions while also dealing with their child’s and likened the grief associated with diagnosis to a bereavement.

> *When your children are very upset, and there’s nothing that you can do, it’s very challenging. I keep using that word. There’s nothing else that I can really say.* (Henry, parent, male)

> *We were plunged into grief. We were just drowning in it*. (Tanya, parent, female)

> *You’re feeling sad and angry as you do going through bereavement.* (Lois, parent, female)

Diagnosis was associated with anxiety, exacerbated by a lack of information about the condition and prognosis.

> *I think I’d definitely be a bit more better off if I’d just known that it wasn’t really that rare and I wasn’t going to go blind and things. If I just knew more about it, then I think I would’ve been a bit less stressed about it.* (Holly, female, age 13-15).

Being diagnosed at a young age was a protective factor for some young participants, as they felt they were too young to fully understand the potential implications of VI, with some reporting their wellbeing being affected more as they grew older:

> *When I first found out about my vision impairment, it was when I was five, maybe, I feel like as I got older, I started to realise how it would affect me as well.* (Claire, female, age 13-15).

> *It’s only in the last year or two, I would say, that really things have started to deteriorate from a mental well-being point of view. From the worries and the anxieties, and the things started to kick in.* (Munira, parent, female)

This was particularly relevant for independent living and travelling:

> She *has a lot of anxiety. When she has to find a taxi, for example, and she uses Uber, they give you a registration number. She can’t see it straight away. That freaks her out. She’s got a fear of being in a taxi and being kidnapped because she’s not sure where she’s going*. (Sue, parent, female)

The impact of VI on wellbeing was described as fluctuating rather than being static, with some days feeling more positive than others.

> *Some days it hits me like a ton of bricks that I can, in fact, not see that, and knowing that it is likely just going to get worse from here*. (Lydia, female, age 16-18).

> *The actual negative feeling tends to only last about a week, and then I get over it. I guess that week is still quite differential or detrimental.* (Mark, male, age 16-18).

Participants discussed these negative impacts as manifesting in different ways including feeling overwhelmed, sad, low in mood, frustrated and angry.

> *A lot of the time, it’s really frustrating having to do this or that just to see a board or something. Obviously, it can be a bit* upsetting. (Holly, female, age 13-15).

Parents were able to distinguish differences in their child’s self-esteem before and after diagnosis, noting that CYP often became more critical of their own abilities following diagnosis, which was associated with lower self-esteem.

> *She has what she describes as her old self and her current self. She feels like her old self was very, very capable. Her current self is a very bright girl, but she feels like the old self had the edge there. She thinks the new self is always tired and her new self is lazy and ineffectual.* (Tanya, parent, female).

Some parents indicated that their children experienced co-occurring mental health conditions, which they linked to the experience of living with a VI or being diagnosed with a serious eye condition.

> *My child is under CAMHS, has been diagnosed with OCD with intrusive thoughts caused by trauma* (Ann, parent, female).

The most extreme impact of VI on well-being was evident in accounts of self-harm and suicidal ideation among CYP, which was reported by parents.

> *He went through a period of self-harming…He would just go into himself and bang his head off a wall out of just frustration* (Lois, parent, female).

> *My child said, “I either went out with my boyfriend to a film I didn’t want to watch, or I stayed here thinking about killing myself.”* (Tanya, parent, female)

Many parents reported that their child did not accept their VI and were in denial regarding the extent of their loss of vision. They linked this lack of acceptance to poor well-being and negative emotions, such as embarrassment, frustration, low confidence, and low self-esteem.

> *He is struggling to accept his visual impairment. I say that because I can see that he’s very embarrassed… Even if I mention or ask whether he can see something, he gets quite frustrated with me and annoyed with me because I’ve mentioned it in public, and he’s worried that other people can hear. He’s very embarrassed by it, which I don’t think is healthy.* (Tanisha, parent, female).

As young people developed more acceptance of their VI, they felt it impacted their well-being less.

> *Hearing that, “Oh, yes, I’m going blind,” doesn’t really affect me now because I know it’s happening. I’m not really bothered about it.* (Mark, male, age 16-18).

> *She’s not desperate to see properly. She’s not looking at every single clinical trial that comes along. She’s just like, “Yes, I’m all right with that, actually,” which is lovely.* (Nicola, parent, female).

> *I think a really good thing is you’ve got to embrace it. You’ve got to embrace that you have the condition. You’ve got to embrace yourself and just see yourself like - A good way to put is, how would you act if you didn’t have the condition? Just do that because you’re still the same person. If we’re talking visual impairment-wise, you’re still the same person, but you’re just seeing a bit less.* (Mark, male, age 16-18).

> *When I think about it, I don’t say to myself, “Oh, I have a vision impairment. I can’t do this, or I can’t do that.” I try to be as optimistic about it.* (Mustafa, male, age 16-18).

Humour was also identified as a means of managing the emotional impact of having a VI, with CYP reflecting that being humorous about their VI helped them manage difficult experiences associated with sight loss. This appeared to serve as a way to alleviate emotional discomfort and provide them with a sense of control over how their VI affected them.

> *Before I turned 18. The sister turned around and went, “On your 18th birthday, do you want to go out and get blind drunk?” She turned around and goes, “Maybe blind drunk wasn’t the best way to explain it. How about we just go out and get you drunk?”* (Lydia, female, age 16-18).

> *Because we’re mates, we actually joke about it a lot, and we just have fun. We just take the piss, to be honest. I don’t care if they make jokes about it because I know I can joke about other things that they do, or all that kind of stuff*. (Mark, male, age 16-18).

### Theme 2. Fighting the system

This theme reflected how parents and CYP navigate both formal and informal support systems, highlighting the barriers they face. It emphasises the advocacy efforts of parents and CYP, while identifying gaps in support provision and the challenges faced in accessing and securing appropriate support.

Parents reported having to constantly advocate for their children. With school and local authorities, this took the form of both efforts to secure access to support and of advocating for others to adjust or reconsider their expectations of the child, often at a high financial and time cost to the parents, which affected their own well-being.

> *If I’m being completely honest. I think the hardest thing is having to fight for his basic rights, which the state puts in place, but despite that, there’s a struggle. I think the hardest thing has been to convince people not to have lower expectations of him. For example, in his studies, we’ve constantly had to convince people that, “Actually, look, he’s really bright, but he needs the right setup, he needs the right access, and he will show his true potential.”* (Tanisha, parent, female).

Young people highlighted the difficulties of advocating for themselves, particularly as young children, and emphasised the importance of their parents advocating on their behalf. However, some CYP described developing the ability to advocate for themselves over time, including both for their own needs and those of the sight loss community.

> *I would say definitely telling their parents what they need because their parents or carers should be able to understand and be able to almost do it for them, especially if they’re quite young. My mom and my dad still do some of the stuff for me because I just don’t really want to do it myself.* (Claire, female, age 13-15).

> *He did the presentation and saying, “I’ve got Stargardt disease, and it means this, and this is what it means.” I think he was quite confident in speaking about it and doing all that*. (Amir, male, parent).

Despite parents being motivated and effective advocates for their children, they identified a lack of awareness of available support as a barrier to accessing assistance. They reflected that it was often not until many years after diagnosis that they became more aware of the provision available, and this was often as a result of their own research or through word-of-mouth within the sight loss community.

> *Nobody reaches out to you and tells you there is A, B, C, D available. You have to seek and you’ve got to really, really search. Years later, we learned that he was eligible for the disability living allowance, I do believe. It’s something that he should have had from day one. We had no idea.* (Munira, female, parent).

> *I’m sure there’s probably more support out there than I’m actually accessing.* (Henry, male, parent).

Several participants emphasised that they were provided with little or no information from the hospital on available support provision at the time of diagnosis. Although some reported receiving limited signposting to services or being provided with a leaflet.

> *There was no leaflet to give us, no mental health support, or what we were to do next, who we should contact. It’s just this floundering about that you end up doing and just hoping that you’ve picked the right thing and you’ve selected the right support.* (Anne, female, parent)

Participants reflected that when support was available, such as information leaflets, it was often designed for elderly adults rather than CYP, which were identified as barriers, as they made the CYP less likely to access the support.

> *When you do find the stuff, the leaflet - for example, the low vision catalogues you get, it’s just old people on it, and it’s really a bit off-putting. If you imagine you’re a 12-year-old kid looking at it and thinking, “I got to use that,” when they saw old people in this booklet using that.* (Anne, Parent, female).

Teachers were identified as sometimes creating barriers to accessing educational support. Participants observed that, often, teaching staff were not competent in using the technology required by the CYP and therefore did not use it.

> *They didn’t sit me at the front, or they didn’t send me the slides that I needed on my laptop, just stuff like that.* (Claire, female, age 13-15).

> *By the end of the first week, she came home in tears because every single lesson that she went into, the teacher was like, “Oh, yes, I forgot that you needed that.” “Oh, I forgot your name so you’re not sat at the front of the classroom.” “Oh, I’ve not printed out your large print stuff, I’m really sorry.” “Oh, I’ve not ordered your maths book with your large squares, so that won’t be here till next week.” “Oh, I don’t know how to use Google Classroom, so I just can’t put the slides on Google Classroom for you.” Every single lesson, she was going to the teacher and going, “I can’t do the work because I can’t see it.”* (Vicky, parent, female).

Participants highlighted poor communication within schools as an additional challenge to their child receiving appropriate support. They explained that primary schools were generally easier to navigate, as fewer staff members interacted with their child, allowing for a more consistent understanding of their needs. In contrast, secondary schools involve multiple teachers, each responsible for many pupils, which can make it easier for individual needs to be overlooked or inconsistently addressed.

> *The communication is not always perfect. You get [a] new teacher, and the teacher would not know that he has to do certain things for people in the class that have certain needs. This is something where the school needs to be maybe a little bit more efficient.* (Matteo, parent, male).

> *School is a massive stressor. School is difficult. School is very, very difficult. Other children with additional needs, I don’t know. There’s just so many kids with additional needs. There’s just so many children. I just don’t think they can cope. You’ve got all the ADHD and autism kids mixed in with all the vision impairment kids, mixed in with the learning difficulty children. There’s just so much going on constantly.* (Lois, parent, female).

The transition from CYP to adult services was also highlighted as a significant challenge, particularly when entering university. Participants observed that once CYP left school, there was much less support available to them, and the support that was available, such as the Disabled Students Allowance, was described as involving a lengthy process, making it difficult for CYP to access independently.

> *Turning 18, you are just forgotten, basically, within the system. Really, I do think that, and I’ve tried to fight it as much as I can. It was just totally forgotten. You’re just a young adult.* (Sue, parent, female).

> *She went to university, we had to apply through the DSA to get all her equipment but that’s the thing in itself. She started September, and it didn’t come till the December. She had to flounder around a bit for the first.* (Anne, parent, female).

### Theme 3. Being seen and being misunderstood

This theme encapsulated how VI influences the way that participants experience relationships, develop a sense of self and navigate their place within society. It reflects how VI can shape interpersonal interactions, such as communication, others’ perceptions and social inclusion, while also affecting how participants understand and construct their own identities. It also explores how participants are positioned within social structures, including the impact of societal attitudes, accessibility, discrimination, stigma and marginalisation. The theme emphasises the ways in which personal experience and broader social contexts shape participants’ lived realities.

Participants identified that having VI could negatively impact interpersonal interactions. This was particularly evident in social situations, with participants describing difficulties recognising people, which left them feeling embarrassed and made it less conducive to making new connections. The effort required to navigate social situations was also reflected upon, with participants often preferring to avoid situations that demanded this additional cognitive and emotional energy. This led participants to feel isolated and lonely.

> *There’s been instances where, for the first time, I said, “Oh, hi, what’s your name?” which is fine. Then I’ve gone back to them again, saying, “Oh, hi, what’s your name?” They’re like, “Oh, you already spoke to me.” I was like, “Oh, sorry, I didn’t realise.” That’s happened before.* (Mark, male, age 16-18).

> *Social situations make her uncomfortable as well. Nightclubbing is not for her because it’s just a sensory overload.* (Sue, parent, female).

Many parents reported that their children experienced difficulties with friendships, which they believed were largely due to VI. Not being able to take part in playground sports and games during breaks was identified as a factor in friendship difficulties, particularly for boys with VI. Some friendship groups were described as tolerant, supportive, and understanding, but parents perceived this acceptance as limited or conditional.

> *Because he doesn’t play ball sports, for example. I think sports, especially with boys, is a big thing.* (Tanisha, parent, female).

> *However good your friends are, they will only sit at the front of the class with you for a certain number of lessons. They won’t sit there all day, every day, unless you’ve got a very, very exceptional friend, and she doesn’t have that very, very exceptional friend yet…Then, of course, it’s not just the fact that she’s in the front on her own, it’s the fact that they’re all at the back talking. Then, when they come out of the lesson, she’s missed out on the whole social interaction. She always feels like she’s a bit behind.* (Vicky, parent, female).

Conversely, CYP participants were mostly positive about their friendships, with friends described typically as being supportive and understanding.

> *My friends know me. If I come into the room, they’ll just call my name out, and so I know where to go. They are very supportive, to be honest. They all understand that I have a visual impairment.* (Mark, male, age 16-18).

> *My friends have adapted. If there’s something, for example, happening at the other side of the lunch hall, they’ll be talking about it and also filling me in on what’s going on.* (Hamza, male, age 13-15).

> *My friends don’t mind.* (Holly, female, age 13-15).

There was also a contrast in how parent participants described the effect of VI on family relationships. Some relationships, particularly between spouses, were reported to be negatively affected, while others were perceived as relatively unchanged or less influenced by VI.

> *I think we focused quite heavy on it and trying to figure out ways of how can we fix this, what can we do? I wouldn’t say we lost our - we kind of derailed slightly. That’s how it went. Now I think about it, being a bit older and going through it, I think that’s just natural. Me and the missus, we felt like we were going nowhere. We had to reconnect again. I think it’s going to do when you find something that can tear you apart a little bit.* (Amir, male, parent).

> *Me and my husband work quite well together. He’s very now in the moment, so he’s living for today, and he was doing all the fun stuff. I was thinking, “In six months, we’ll go traveling so my child can see all these amazing things.” I was thinking really far ahead, and he was thinking in the now. Between us, we sorted it out.* (Anne, female, parent).

Differences emerged in how the CYP participants viewed the impact of VI on family relationships when compared to their parents. They generally perceived family relationships as very supportive and unaffected by VI, which may suggest that parents of CYP with VI are adept at masking challenges and reducing their impact on their children.

> *At home, as long as you have a family that helps you and understands you and tries to get things for you that you need…it helps.* (Mustafa, male, age 16-18).

Participants highlighted that VI is often a hidden disability. They reflected that non-visually impaired people are often either completely unaware that they have VI, or they will consider them to be exaggerating the extent of their VI.

> *It is like an invisible disease. You wouldn’t know with my child, it’s not apparent at all. They said people often assume that you are either lying about it, or you are over-egging it, or you’re making a big deal about something that’s like-- so, and I think, unfortunately, that does stop people when they do need help.* (Nicola, female, parent).

> *I think people need to be a bit more just aware that there are conditions that you can’t really see much*. (Holly, female, age 13-15).

Participants’ accounts highlighted how VI being a hidden disability shaped social interactions. While some participants reported that they were comfortable with others’ curiosity about their condition and understood why others might be curious, others found it draining and difficult to keep explaining the extent of their VI.

> *I honestly don’t mind if they come and ask me because I would do the same, honestly.* (Claire, female, age 13-15).

> *To walk into an environment of people who you’ve never met, who are able-bodied, and try to tell them that you’ve got a vision impairment is not easy.* (Sue, female, parent).

Participants shared experiences of bullying associated with VI, which they highlighted was often driven by others’ lack of understanding or disbelief, particularly where the impairment was not immediately visible.

> *All the ones that I grew up with decided that I was faking it because I had friends that were visually-impaired, so decided they were going to start bullying me, which wasn’t the best time.* (Lydia, female, age 16-18).

Experiences of bullying did not relate to isolated behaviours. It was reported that negative peer behaviours were underpinned by wider discriminatory attitudes within society, including disbelief and a lack of understanding about VI. This led to some participants being excluded from activities and sports.

> *Even his QTVI [Qualified Teacher for Vision Impairment] to be honest, it’s very, “Oh, well, never mind,” sort of attitude if he doesn’t get to do sports, or, “We’ve done all we can. You can’t really do much about it. It’s a good school.” They’re very ready to accept that he doesn’t do certain things that his peers would do.* (Tanisha, female, parent).

> *The first club I had to quit because they were like, “Oh, just do this,” but like, “She can’t see that,” and then they wouldn’t give anything for alternative.* (Lydia, female, age 16-18).

Socialising with other CYP with VI was identified by many CYP participants as an important way of normalising experiences and reducing the burden associated with VI as a hidden disability. This differs from socialising with peers without VI, where participants were often required to explain or justify their condition; interactions with peers who also had VI provided a shared understanding that removed the need for repeated explanations.

> *Meeting other people with the same thing, because I think that really helps to put it into perspective and realise it’s not only you and things.* (Holly, female, age 13-15).

> *It’s nice to catch up with them, seeing them again, and then know that we’ve got a good bond and dynamic going on. For me at least, that helps my head. It is a completely different thing from school and work because I don’t go to any other group, so it’s really nice to be a part of it.* (Mark, male, age 16-18).

> *I think connecting them to communities or parents or kids of the same age, I think, really helps. Because it’s real-life experience. You can go to someone who works in that environment and has seen those cases. When you’re actually living it and talking about it, it’s different. I think that gives you a bit more confidence that you’re hearing something a bit more real.* (Amir, male, parent).

However, some parents expressed hesitation about their children socialising with other CYP with VI, fearing that involvement might lead to their child being negatively labelled through association with other people with sight loss or that it could prompt difficult emotional responses by requiring them to confront the realities of living with VI.

> *She doesn’t see herself as that’s the main thing about her. I think she’s happy to talk to people, obviously, with the same condition, but she doesn’t feel a need to be part of a community about it.* (Nicola, female, parent).

Social media was noted as a way for some CYP with VI to make social connections and reinforce a sense of shared experience and belonging.

> *She’s just met some great people of all ages. I do know that she has contact with them through social media, which I think is really important. She checks in with people online.* (Tanya, female, parent).

> *I know she follows a couple of people on Instagram and things like that with Stargardt’s and visual impairment.* (Nicola, female, parent).

Living independently was linked with wellbeing and how participants were perceived by others, particularly in relation to their independence. Some were described as highly independent young people, managing day-to-day life in ways comparable to their non-visually impaired peers.

> *At a very young age, we’d go into London on the train and show him the trains, show him this, show him that. Now he’s confident, he can just go anywhere he wants. He’ll just use the train, use the buses, or wherever. We worry all the time, but we’ve got to let him go, and he’s confident in those things.* (Amir, male, parent).

> *He’s been in charge pretty much of managing his own condition by himself and being responsible towards that.* (Matteo, male, parent).

Others, however, were viewed as more highly reliant upon family support, reflecting the additional practical and environmental challenges they face. Parent participants reflected on the level of support they provide their children, noting that much of it has become so embedded in daily routines that it goes unnoticed. Additionally, they recognised the substantial barriers their children encounter when striving for greater independence.

> *I guess there’s times when we don’t really realise the support that we give her because it’s part of our day-to-day life. Whereas, I suppose, when we go and see friends, and we see the things that we do for our child that they don’t do for their children, then if we think about it, we notice it.* (Vicky, female, parent).

The challenge between encouraging independence and managing risk was identified as leading to feelings of frustration for both CYP and parents, particularly when efforts to be more independent are thwarted. However, these experiences highlighted the determination of CYP participants to build autonomy and their families’ efforts to support this.

> *For a child who is extremely independent in a lot of ways, is also very needy in a lot of ways.* (Sue, female, parent).

> *At Christmas, she screamed at me and went, “Do you think I effing like being codependent on you? I bloody don’t.”* (Tanya, female, parent).

### Theme 4. Building a life with vision impairment

This theme centres on how participants function in everyday contexts whilst living with VI, alongside their aspirations for building their lives and living well in adulthood. The theme focuses on adaptation to challenges, participation in daily activities, and future-making as CYP with VI begin to shape their pathways into adulthood, with a focus on building fulfilling and independent lives.

Within this context, participants reflected on their experiences of counselling as one way of adapting to the emotional challenges associated with VI. Many described positive experiences of counselling and the benefits that it brought. CYP participants highlighted how counselling supported them to build a more positive mindset, come to terms with their VI and manage some of the difficult emotions they experienced. In doing so, counselling was seen as contributing not only to day-to-day well-being but also to longer-term confidence and capacity to engage in education, relationships and future planning.

> *I’ve had counsellors before. I guess they really helped as well, especially at a young age. I think that those things have really helped in my head to keep me having an ongoing good mindset, a positive mindset.* (Mark, male, age 16-18).

> *The therapist I was given was really good. She was good.* (Holly, female, age 13-15).

Parents also discussed the benefits of counselling for their children; however, their views were more nuanced than those of the CYP, recognising that counselling was part of a larger support strategy, with the impact difficult to quantify from anecdotal evidence alone.

> *She does have weekly counselling with the hospital, which I think is very helpful. I think the counselling has been very good.* (Henry, male, parent).

> *The school arranged for him to have counselling to help him better express his anger and frustration… Has it helped? It’s not harmed him definitely. I would always take any help that is on offer. I don’t know. It’s difficult to quantify it. It’s difficult to quantify it because there are so many moving parts.* (Lois, female, parent).

Despite parents and CYP participants highlighting the benefits of counselling, some parents felt that it had not been beneficial for their child.

> *He received some support out of XXX, but then the funding for that ended, and then he was eventually transferred to CAMHS [Child and Adolescent Mental Health Services]. Then I think it just didn’t make any difference, and my son wasn’t very happy, so we just left it.* (Tanisha, female, parent).

> *He’s not overly good with being face-to-face with new people. They put him in a room with the counsellor and left him, and he didn’t really talk. Then the next time, it was the same again. I was like, “It’s not working.”* (Munira, female, parent).

Some parents reported that they had sought and received counselling support themselves to help them cope with the challenges of having a child who was living with VI, which was beneficial.

> *I just had phone counselling weekly with them, and that absolutely helped me so much just to come to terms with it and sort of look at it a bit more brightly, and that was just completely invaluable. They were brilliant.* (Nicola, female, parent).

> *I’ve accessed support, and we were lucky that we got on. I think there’s a parents’ course that you can do with XXX. We did that for six weeks.* (Anne, female, parent).

Whilst participants’ reports varied with respect to how helpful they had found counselling, there was a clear consensus that CYP with VI should have access to free counselling. Participants emphasised that this support should be delivered by clinicians with an understanding of sight loss and, ideally, be specifically tailored to the needs and experiences of CYP with sight loss. Parents highlighted the importance of offering counselling as an early intervention, to enable timely support as CYP begin to adjust to their VI.

> *Counselling, the space to be able to talk about how they’re feeling without the risk of worrying somebody that they’re close to or so they can just really be honest about it and explore their own feelings, I think is really key. That’s a really important thing if kids can use it.* (Fran, female, parent).

> *Definitely near the start, because it was really weird realising that you can’t really see anything anymore. I think that’s probably what people need the most help with.* (Holly, female, age 13-15).

Participants reported that assistive technology is a key resource in supporting adaptation to VI by reducing barriers, enabling engagement in schoolwork, hobbies and social life, aiding independence and self-efficacy and increasing confidence.

> *She has an iPad that’s Bluetooth to the whiteboard so she can see what’s going on*. (Nicola, female, parent).

> *We have ordered some glasses to read. They’re called the Meta Ray-Ban. They have a feature that called Look and Tell, so it can read things if I ask, “What does that say?” It can describe things as a camera, so it can do that. We’ve ordered that and I think that can impact my life positively.* (Hamza, male, age 13-15).

> *They’re called Ray-Ban Meta glasses, and they have a camera inside them. I can ask them what I’m looking at. I use it at restaurants menus. I use it in menus in restaurants, and I use it on food packaging or in the train station.* (Mark, male, age 16-18).

However, participants noted that the price of assistive technology can be prohibitive and therefore not accessible to all CYP with VI, and that using assistive technology can, at times, accentuate feelings of difference from peers.

> *She isn’t taking her big laptop with her because she’s so terrified of somebody sitting behind her and seeing her magnified text. Even though we’ve put lots of things in place, I don’t think she’s accessing a lot of them because she doesn’t want to be outed and judged.* (Tanya, female, parent).

> *I didn’t really have the luxury of cycling through tech to explore what was best for me.* (Lydia, female, age 16-18).

Mobility training was recognised as an important form of support, assisting CYP to adapt to the practical challenges of VI, reduce reliance on others, increase access to school, work and social activities and develop the independence required for adulthood. Participants described access to mobility training as timely and sufficient, with no gap in provision identified.

> *Habilitation is available. That was really good. I thought that was really useful. The fact that habilitation from the council is available. We had really good access to it, and our council did a particularly good job.* (Munira, female, parent).

> *They sent someone out, and that person spent time teaching my child how to walk backwards and forth to school safely. They went into the school when she was struggling to get around the school. They spent a lot of time doing techniques and ways in which she could walk around the school safely.* (Vicky, female, parent).

Participants described taking part in many different hobbies and sport which were identified as important aspects of participation in everyday life. Not only were these activities seen as providing opportunities for enjoyment, social connection and the development of confidence and identity, but they also support day-to-day well-being and foster a sense of belonging.

> *She does Guides. She does the piano. She’s done various clubs throughout her time and they’ve all been very-- she’s gone on the Guide camp and things, and that’s all fine. She loves music and she loves playing the piano.* (Vicky, female, parent).

> *I really like drawing. That’s really the main thing. Art, textiles*. (Holly, female, age 13-15).

> *She’s got a very strong hobby, which has kept her going throughout the years… It’s the one thing she can do which makes her the same as everybody else. I think that’s the reason. That element keeps her going, if that makes sense.* (Sue, female, parent).

However, access to sport and hobbies was sometimes described as difficult, with some participants facing barriers that required adaptation, limited their involvement, or prevented them from participating completely.

> *He can’t play rugby or football because he can’t see the ball coming. He can’t take part in a team sport because it’s not going to be fair on the rest of the team. How are they going to make a reasonable adjustment? If they compete, the logistics of it is just too difficult. It’s not fair on the other people.* (Lois, female, parent).

> *He likes cycling, things like that, individual sports which don’t involve people running around and going beyond his line of sight. He doesn’t have the opportunity to do that. We have tried outside school as well, but it’s just really tricky to set that in place.* (Tanisha, female, parent).

Support animals, such as buddy dogs, were described as assisting participation in everyday life by increasing confidence in social situations and reducing reliance on others. They were also observed to improve well-being and support the mental health of the CYP. However, support animals such as buddy dogs involve taking on responsibility for the animal, and access to this form of support is not widely available. The cost of such support was also a limiting factor, and such costs could be prohibitive for many families.

> *We’re also getting a buddy dog. We’ve been working with the Guide Dogs…He will often ask me, “Oh, is this going to be my best friend? Is this my dog? Is he there to help me?” I can see that he’s already viewing this dog as a support tool. I can see that this is really important for him.* (Lois, female, parent).

> *I knew horses have actually got a really amazing effect. I managed to find a lady locally who did equine therapy. She went to her for probably 18 months. She would just go hang around with the horses. Sometimes they would just chat and stroke them, sometimes they’d be a bit of riding. Sometimes they were all asleep. Her and the lady would just sit in the sunshine next to snoring horses. It was part therapy, part giving you a break away from your studies. That worked really, really well. It was getting a little bit expensive for us as a family.* (Tanya, female, parent).

Parents reflected on how VI impacted their child’s career aspirations and choices. For some CYP, this involved adapting their career goals in response to perceived barriers. These considerations were linked to their experiences of participation in education and daily life, which shaped confidence and expectations for the future.

> *She was going to go and study physiotherapy. I think she realised or didn’t want another three years of studying with having to have more support than her peers, really.* (Sue, female, parent).

However, the CYP demonstrated determination in pursuing their career goals and were able to study at university or be employed if employers understood their condition and made suitable adaptations.

> *The only thing that I don’t do is work on the till because I can’t really see the till. The other thing is that we have these printed out receipts for what’s been ordered. I can’t read them, but I just get my colleagues to read them out for me. I’m really close with all the colleagues, so it works out really well because they understand my condition and they understand how it affects me.* (Mark, male, age 16-18).

## DISCUSSION

Vision impairment has emotional, social, and systemic consequences, consistent with Ecological Systems Theory.[32,33] Both the experience of the young person with VI and the environment around them will affect their adaptation to sight loss and their wellbeing. We identified low acceptance of vision loss; low functional adaptation; reduced self-efficacy; effects of social stigma and discrimination as factors that linked VI to reduced mental wellbeing. Supportive strategies described by participants as supporting their mental wellbeing included social connection and counselling. Internal support strategies such as a sense of humour, artistic and creative endeavours were also identified as beneficial.

This work adds to our previous qualitative research with adults with VI, which identified emotional, practical, medical and societal impacts of low vision, with self-support and external support strategies helping their wellbeing.[12] We used a similar approach to Roborel de Climens and colleagues who also interviewed young people with inherited macular disease and their parents, although their focus was on practical limitations on daily life which medical or surgical treatment could theoretically benefit rather than on mental wellbeing.[34] Despite not asking questions about wellbeing specifically, these researchers did identify that people with VI experienced frustration, annoyance, irritability, exasperation, being moody and anger.

A strength of our study was the focussed recruitment strategy of focusing only on a single cause of VI – inherited macular disease – rather than a more diverse group of adolescents with sight loss of different aetiology, such as cerebral VI which may affect people in a very different way. We were particularly interested in those who developed VI in childhood or adolescence rather than those who were vision impaired from birth. Interviewing parents or carers of these young people provided additional validation of the themes identified. Of course, this limits the applicability of our research to people with different forms of VI.

A further limitation is that all of our participants were recruited from a specialist, tertiary level ophthalmology centre and were under the care of medical, rehabilitation and support services, so may therefore not represent the broader population of people with VI. In particular, the impact of VI on young people who are not under this level of medical care, or who are underserved by health services for reasons of social exclusion, socioeconomic status or migration status should be explored. As participation in the study was voluntary, people with very low levels of wellbeing may have chosen not to participate, leading to potential selection bias. However, parents of these people may have been *more* keen to take part in this research.

Our findings suggest that targeted support interventions designed to address and overcome these challenges should be provided at an early stage and by therapists experienced in VI. To address difficulties in information at the point of diagnosis, the support of an age-appropriate Eye Clinic Liaison Officer should be provided, and comprehensive information leaflets should be supplied in accessible and adolescent-friendly format. This may reduce the likelihood of people developing post-visual diagnosis distress,[37] and speed their adaptation to VI. Peer support should be offered to ensure higher levels of participation and lower levels of loneliness,[9] especially given the link between loneliness and wellbeing in people with VI,[38] although the efficacy of peer support is not clear.[19] Specific training in VI for teachers and other staff working with young people may help with interactions between people with VI and their non-vision impaired peers. The importance of support during transitional phases between different educational settings (for example, school to university) or between education and the workplaces was emphasised by our participants. ACT has been suggested as a mechanism for supporting young people with VI and has been shown to be effective in a small sample.[24] The emotional impact of VI on parents and carers was also clear throughout our interviews, so support should be provided for caregivers of young people with VI.[39] Future research should examine the impact of a multidisciplinary approach of emotional and practical support for young people with VI.

## Data Availability

Anonymised data from the present study are available upon reasonable request to the authors.

## SUPPLEMENTAL MATERIAL: INTERVIEW TOPIC GUIDES

*Interview topic guide for CYP. (VI=visual impairment)*.

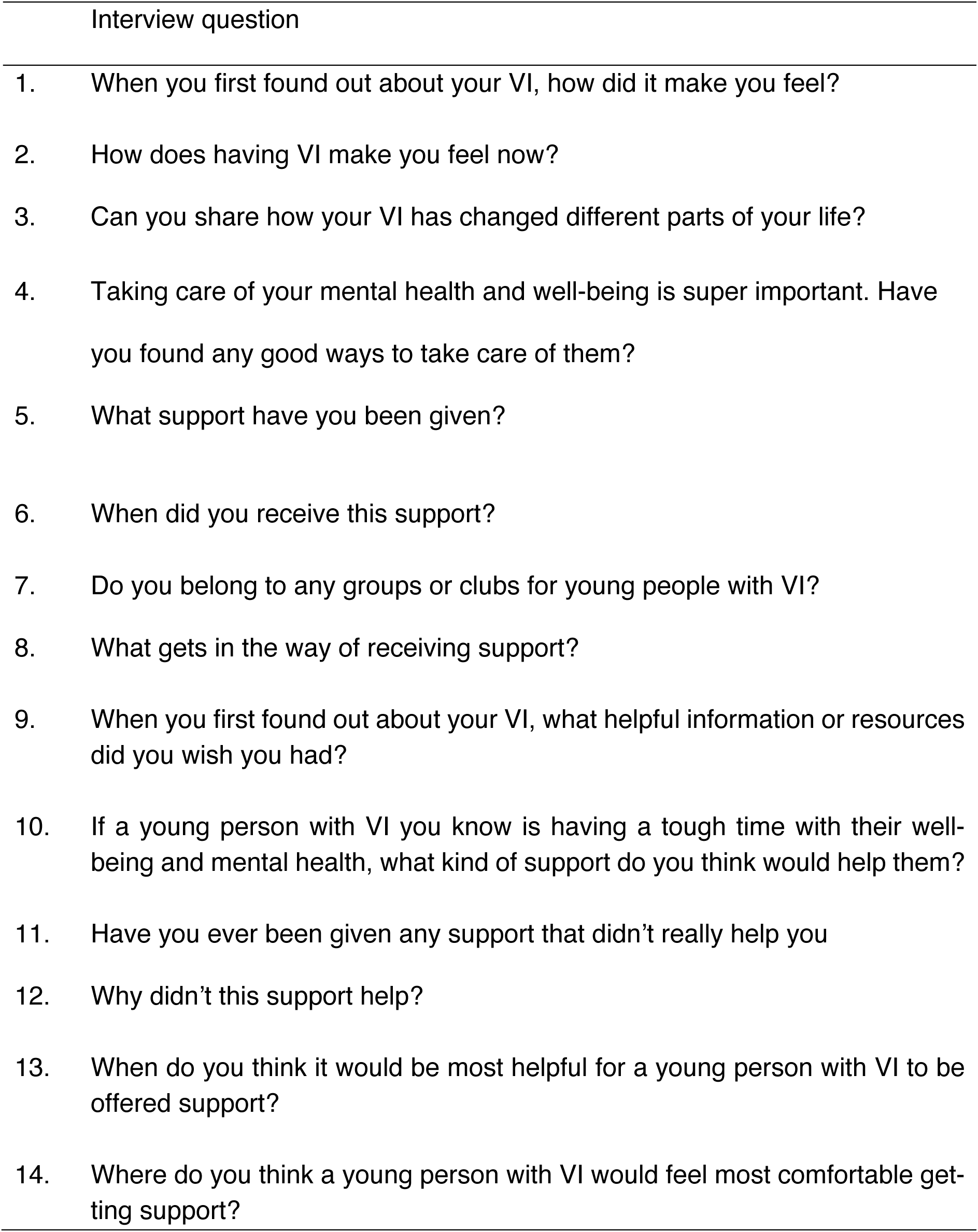

*Interview topic guide for parents/carers. (VI=visual impairment)*.

|  | Interview question |
| --- | --- |
| 1. | How would you describe your child's well-being when they were first told they had this eye condition? |
| 2. | How would you describe your child's well-being at the current time? |
| 3. | How would you describe your well-being? |
| 4. | What support does your child currently receive? |
| 5. | Are they members of any groups of people with VI, or do they know anyone else with VI? |
| 6. | Are you a member of any VI groups? Do you know any other parents of a child with VI? |
| 7. | What else would have helped your child/ you/ your family that you haven't had? |
| 8. | Is there any support you've had which wasn't helpful? |
| 9. | What support do you think we should offer to young people with VI who are struggling with their mental health and well-being? |
| 10. | What are the biggest hurdles to receiving support? |
| 11. | Where is the best place to give this support? |
| 12. | What do you think makes your child's mental health and well-being better? |
| 13. | What do you think makes your child's mental health and well-being worse? |
| 14. | As a family what is the most helpful thing you do for your mental health and well-being? |

## SUPPLEMENTARY MATERIAL: COREQ CHECKLIST

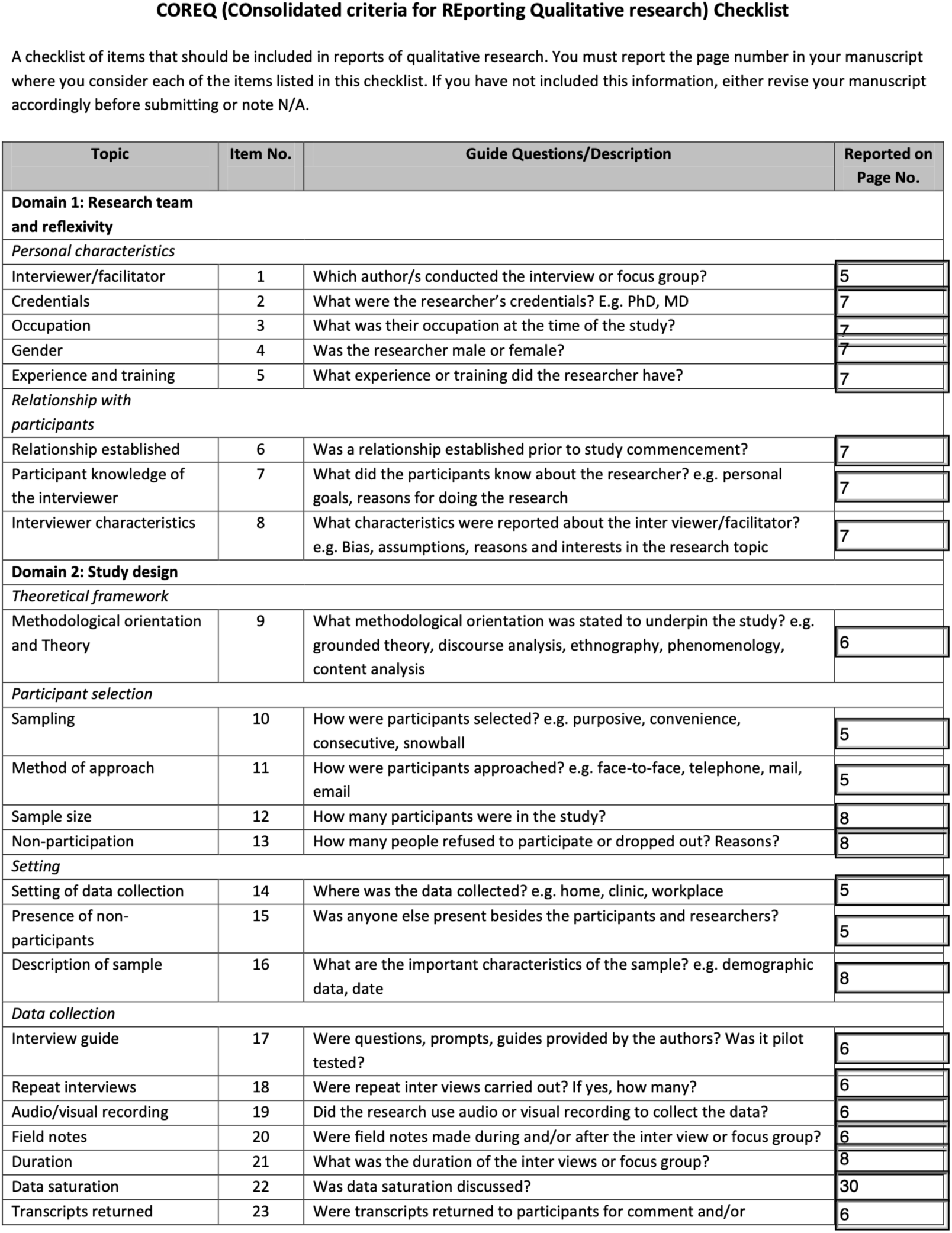
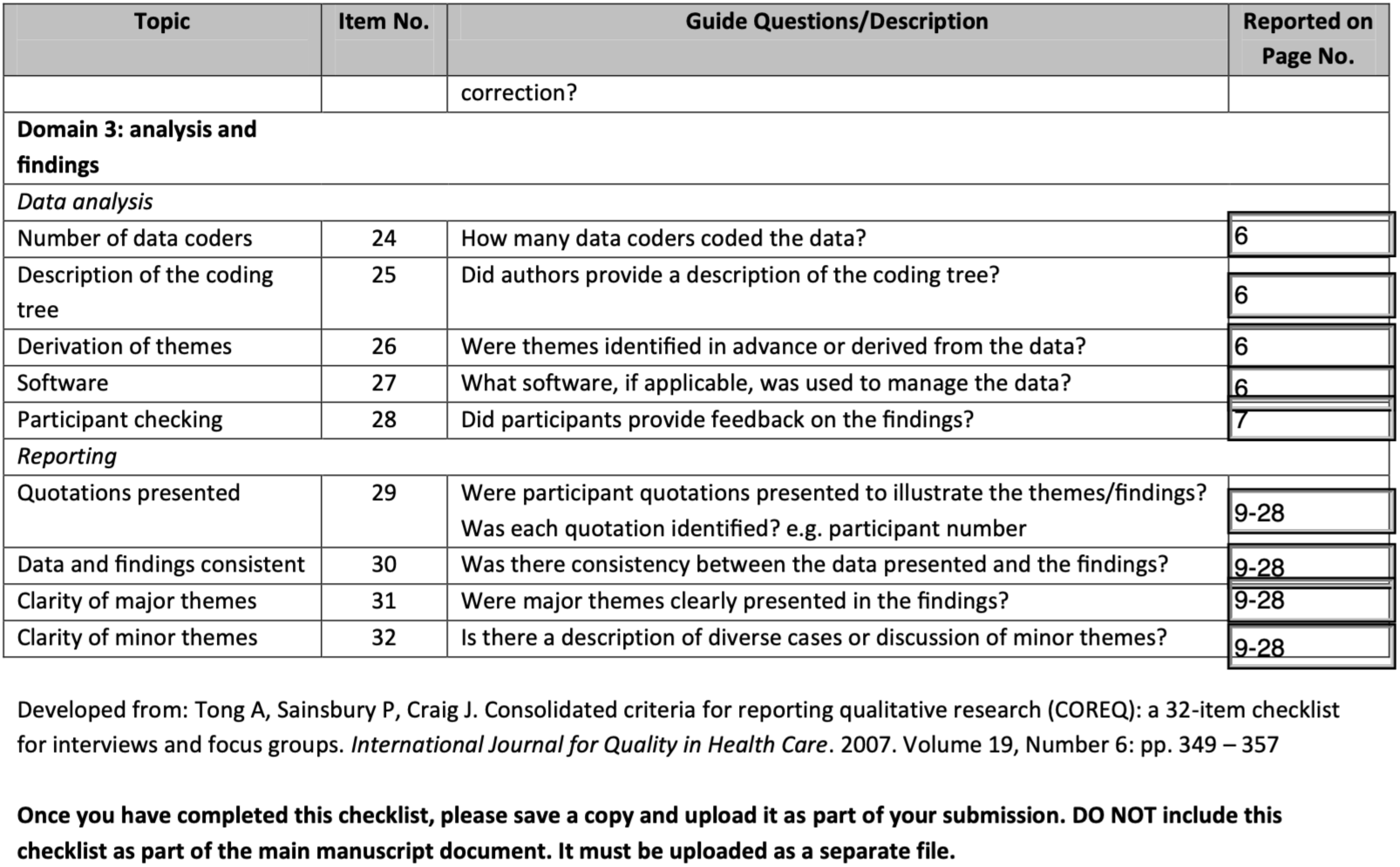

| Topic | Item No. | Guide Questions/Description | Reported on Page No. |
| --- | --- | --- | --- |
| **Domain 1: Research team and reflexivity** |  |  |  |
| *Personal characteristics* |  |  |  |
| Interviewer/facilitator | 1 | Which author/s conducted the interview or focus group? | 5 |
| Credentials | 2 | What were the researcher's credentials? E.g. PhD, MD | 7 |
| Occupation | 3 | What was their occupation at the time of the study? | 7 |
| Gender | 4 | Was the researcher male or female? | 7 |
| Experience and training | 5 | What experience or training did the researcher have? | 7 |
| *Relationship with participants* |  |  |  |
| Relationship established | 6 | Was a relationship established prior to study commencement? | 7 |
| Participant knowledge of the interviewer | 7 | What did the participants know about the researcher? e.g. personal goals, reasons for doing the research | 7 |
| Interviewer characteristics | 8 | What characteristics were reported about the inter viewer/facilitator? e.g. Bias, assumptions, reasons and interests in the research topic | 7 |
| **Domain 2: Study design** |  |  |  |
| *Theoretical framework* |  |  |  |
| Methodological orientation and Theory | 9 | What methodological orientation was stated to underpin the study? e.g. grounded theory, discourse analysis, ethnography, phenomenology, content analysis | 6 |
| *Participant selection* |  |  |  |
| Sampling | 10 | How were participants selected? e.g. purposive, convenience, consecutive, snowball | 5 |
| Method of approach | 11 | How were participants approached? e.g. face-to-face, telephone, mail, email | 5 |
| Sample size | 12 | How many participants were in the study? | 8 |
| Non-participation | 13 | How many people refused to participate or dropped out? Reasons? | 8 |
| *Setting* |  |  |  |
| Setting of data collection | 14 | Where was the data collected? e.g. home, clinic, workplace | 5 |
| Presence of non-participants | 15 | Was anyone else present besides the participants and researchers? | 5 |
| Description of sample | 16 | What are the important characteristics of the sample? e.g. demographic data, date | 8 |
| *Data collection* |  |  |  |
| Interview guide | 17 | Were questions, prompts, guides provided by the authors? Was it pilot tested? | 6 |
| Repeat interviews | 18 | Were repeat inter views carried out? If yes, how many? | 6 |
| Audio/visual recording | 19 | Did the research use audio or visual recording to collect the data? | 6 |
| Field notes | 20 | Were field notes made during and/or after the inter view or focus group? | 6 |
| Duration | 21 | What was the duration of the inter views or focus group? | 8 |
| Data saturation | 22 | Was data saturation discussed? | 30 |
| Transcripts returned | 23 | Were transcripts returned to participants for comment and/or correction? | 6 |
| **Domain 3: analysis and findings** |  |  |  |
| *Data analysis* |  |  |  |
| Number of data coders | 24 | How many data coders coded the data? | 6 |
| Description of the coding tree | 25 | Did authors provide a description of the coding tree? | 6 |
| Derivation of themes | 26 | Were themes identified in advance or derived from the data? | 6 |
| Software | 27 | What software, if applicable, was used to manage the data? | 6 |
| Participant checking | 28 | Did participants provide feedback on the findings? | 7 |
| *Reporting* |  |  |  |
| Quotations presented | 29 | Were participant quotations presented to illustrate the themes/findings?<br>Was each quotation identified? e.g. participant number | 9-28 |
| Data and findings consistent | 30 | Was there consistency between the data presented and the findings? | 9-28 |
| Clarity of major themes | 31 | Were major themes clearly presented in the findings? | 9-28 |
| Clarity of minor themes | 32 | Is there a description of diverse cases or discussion of minor themes? | 9-28 |

Developed from: Tong A, Sainsbury P, Craig J. Consolidated criteria for reporting qualitative research (COREQ): a 32-item checklist for interviews and focus groups. *International Journal for Quality in Health Care*. 2007. Volume 19, Number 6: pp. 349 – 357
**Once you have completed this checklist, please save a copy and upload it as part of your submission. DO NOT include this checklist as part of the main manuscript document. It must be uploaded as a separate file.**

